# LONGITUDINAL OVARIAN RESERVE CHANGES IN WOMEN WITH BREAST CANCER RECEIVING ADJUVANT CHEMOTHERAPY OR TAMOXIFEN-ALONE

**DOI:** 10.1101/2020.06.21.20136689

**Authors:** Shari B Goldfarb, Volkan Turan, Giuliano Bedoschi, Enes Taylan, Nadia Abdo, Tessa Cigler, Heejung Bang, Sujita Patil, Maura N Dickler, Kutluk H Oktay

## Abstract

**Background:** To determine the longitudinal impact of adjuvant chemotherapy and tamoxifen-only treatments on ovarian reserve by serum anti-Mullerian hormone (AMH) levels in women with breast cancer.

**Methods:** One-hundred-and-forty-two women with a primary diagnosis of breast cancer were prospectively followed with serum AMH assessments before the initiation, and 12, 18 and 24 months after the completion of adjuvant chemotherapy or the start of tamoxifen-only treatment. The chemotherapy regimens were classified into Anthracycline-Cyclophosphamide-based (AC-based) and Cyclophosphamide-Methotrexate+5-Fluorouracil (CMF). Longitudinal data were analyzed by mixed effects model for treatment effects over time, adjusting for baseline age and BMI.

**Results:** Both chemotherapy regimens resulted in significant decline in ovarian reserve compared to the tamoxifen-only treatment (p<0.0001 either regimen vs. tamoxifen for overall trend). AMH levels sharply declined at 12 months but did not show a significant recovery from 12 to 18 and 18 to 24 months after the completion of AC-based or CMF regimens. The degree of decline did not differ between the two chemotherapy groups (p=0.53). In contrast, tamoxifen-only treatment did not significantly alter the age-adjusted serum AMH levels over the 24-month follow up. Likewise, the use of adjuvant tamoxifen following AC-based regimens did not affect AMH recovery.

**Conclusions:** Both AC-based and CMF regimens significantly compromise ovarian reserve, which does not recover during the 12-24-month post-chemotherapy follow up. In contrast, tamoxifen treatment does not seem to alter ovarian reserve. This novel information should be valuable for fertility preservation counselling and in assessing future reproductive potential of breast cancer survivors.

## Introduction

Breast cancer is the most common malignancy among premenopausal women, with more than 25,000 cases diagnosed annually in women under 45 years of age in the US alone (1,2). As societal shifts have led to increased rates of women delaying childbearing, a diagnosis of breast cancer is increasingly more likely to occur prior to the completion of family building. Recent advancements in systemic therapies have drastically improved 5-year relative survival rates, which now approach 90% (2). This improvement in prognosis has allowed providers to dedicate more attention to optimizing patient quality of life, including reproductive function. Chemotherapy remains the backbone of multimodality therapy for many young breast cancer patients; however, its indisputable survival benefits often come at the expense of fertility. By inducing DNA damage and apoptosis in primordial follicles and potential microvascular and stromal damage in the ovary, cytotoxic chemotherapeutic agents deplete ovarian reserve and compromise ovarian function and fecundability (3-5). Different chemotherapy regimens confer varying degrees of gonadotoxicity with concomitant rates of decline in ovarian function. The induction of amenorrhea appear to be related to chemotherapy dose and patient age (6). Additionally, with impaired DNA double stranded break repair mechanisms, BRCA carriers are particularly vulnerable to the gonadotoxic effects of chemotherapy and may experience even more profound declines in ovarian function (7,8).

Though menstrual history has been used as a surrogate for reproductive potential with evidence of improved fecundability in patients who resume menses following cessation of chemotherapy, the ability to estimate the extent of chemotherapy-induced gonadotoxicity with such indirect surrogate for any given patient remains elusive. Quantifying reproductive potential remains a challenge with a growing body of evidence supporting the use of anti-Mullerian hormone (AMH) as a biomarker of ovarian reserve (9-14). A glycoprotein and member of the transforming growth factor-B superfamily, AMH is secreted exclusively by granulosa cells of preantral and early antral follicles (15,16). AMH has been implicated in the modulation of ovarian follicle development, both presumably suppressing recruitment of primordial follicles as well as attenuating antral follicle response to follicle-stimulating hormone to establish a dominant follicle (17,18). The population of developing follicles reflects the size of the primordial follicle population, thus designating AMH an indirect marker of the primordial follicle reserve and ovarian function (12).

AMH has been used as a predictor for response to ovarian stimulation, as well as a marker of reproductive lifespan (19-24). It has also emerged as a tool to predict development of chemotherapy-induced amenorrhea and estimate reproductive potential after chemotherapy (25-29). Declining AMH levels during chemotherapy administration coupled with sustained depressions following completion of chemotherapy have been previously observed in short-term studies, reflecting depletion of the primordial follicle pool (25,27-29).

However, there is a paucity of longitudinal data defining the long-term trends in AMH levels in breast cancer patients treated with chemotherapy. To be able to quantify the potential impact of chemotherapy administration on an individual patient’s reproductive potential would be an invaluable tool to guide accurate patient counseling and decision-making regarding fertility preservation (30). As chemotherapeutic agents damage both primordial follicles and AMH producing developing follicles, AMH levels show a sharp decline immediately after treatments. However, if there is remaining primordial follicles, they will give rise to newly developing follicles which will eventually produce AMH, resulting in a “recovery” in levels. Understanding these recovery patterns will also help counsel women and couples on their fertility potential post chemotherapy. Furthermore, the impact of tamoxifen treatment on serum AMH levels is also unknown. Though tamoxifen is not a cytotoxic drug, it is an ovarian-stimulant and can alter ovarian follicle dynamics and hence serum AMH levels.

Therefore our a priori hypothesis was the anthracycline and cyclophosphamide based (AC-based) and CMF regimens but not the tamoxifen-alone treatment will result in significant decline in ovarian reserve. Secondarily, based on the ovarian follicle and AMH production physiology, we hypothesized that the major and clinically meaningful recovery of ovarian reserve from chemotherapy would be at 12 months post-chemotherapy. To that end, we report AMH levels in premenopausal breast cancer patients before, during, and after treatment with chemotherapy or tamoxifen-alone.

## Methods

### Patient selection

This study was approved by the Institutional Review Boards at all participating institutions (*clinicaltrials*.*gov* identifier NCT00823654). Enrollment began in January 2009, as part of an NIH-funded translational research project (NICHD and NCI; RO1 HD 053112) to assess the impact of breast cancer chemotherapy on ovarian reserve and ended in November 2017. Women who had prior chemotherapy or ovarian surgery and those who did not have regular periods were excluded. We also excluded those that were age 45 or older as they may be approaching menopause during follow-up and women who had a recurrence and needed additional chemotherapy.. Participants provided blood samples prior to chemotherapy and at 12-, 18- and 24-months post treatment.

### Specifics of the serum AMH analysis and the assay

Resulting sera were aliquoted and stored first at −80°C then long term at −273°C. Frozen aliquots were transported on dry ice to Webster, TX, where serum AMH was measured on site at Ansh Laboratories by one of the team members using a two-site enzyme-linked immunoassay (picoAMH ELISA, Ansh Labs, Webster, TX) following the manufacturer’s instructions. AMH levels were expressed in ng/mL and all assays were performed within three days in mid-August 2017 with a single lot of reagents. Samples were initially diluted 1:10. The reportable range was 0.003 to 23 ng/mL. Initial values falling below 0.03 ng/mL were retested without dilution and any samples with initial values >11.5 ng/mL were retested at a 1:20 dilution. All final optical densities fell within the standard curve. The coefficient of variation for the four-levels of pooled serum quality controls tested along with study specimens were all < 7%.

### Statistical Analyses

A priori power calculations originally done in study planning were made based on the assumption that there would be at least 0.18 ng/ reduction in serum AMH levels at the 12-month time point with 80% power and type I error of .01 (Type I error is <0.05 to account for multiple comparisons) compared to baseline with repeated measures of ANOVA. Based on the distribution of treatments at the time of the study planning, we anticipated >50% of women to receive AC-based treatments with CMF and tamoxifen-only being evenly distributed. Based on these assumptions, we calculated a sample size of 190 women with breast cancer with about 150 completing follow up and remain evaluable.

A mixed effects regression model was fitted with time, treatment group and time-and-treatment interaction for longitudinal data, adjusting baseline age and body mass index that can influence AMH levels, where random intercept was used to capture within-person correlation (31). Regression and inference were performed on the logarithmically transformed (log10) AMH data, while untransformed (raw) data were used for summary statistics and graphs. AMH levels are dependent on the woman’s age. A (univariate) generalized linear model was fitted on the referent population (e.g., tamoxifen group with baseline data only) to model AMH levels as a function of age and to derive ‘expected’ value of AMH at given age. Statistical analyses were performed with SAS 9.4 (SAS Institute, Cary NC).

## Results

### Description of the study population and the correlation of baseline AMH with age

Out of the 207 women with breast cancer diagnosis who were enrolled initially, 30 were excluded later because they lacked a baseline AMH and 22 were excluded because they lacked a follow up AMH assessment. Of the 155 women, 13 were excluded for receiving other chemotherapy regimens (cyclophosphamide and doxetaxel n=8; vinorelbine, n=1; trastuzumab and paclitaxel; n=4). Of the 142 remaining evaluable women, 106 received AC-based regimens: doxorubicin, cyclophosphamide and paclitaxel (n=95); epirubicin, cyclophosphamide and paclitaxel (n=4); doxorubicin, cyclophosphamide and eribulin (n=4); doxorubicin, cyclophosphamide and nab=paclitaxel (n=2); doxorubicin, cyclophosphamide and lapatinib (n=1). Of the remaining, 19 received CMF (Cyclophosphamide, Methotrexate, 5-Florouracil) while 17 received tamoxifen therapy alone. Characteristics of those 142 patients are summarized in Table 1. Of those, 80 women completed all three follow ups, 126 completed at least two and 142 completed at least one follow up (Figure 1). The most frequently completed follow up was at the 12-month time point (127 women). Although women with no follow up AMH levels were excluded from the longitudinal analysis, baseline AMH values from 5 such excluded participants (so total 17+5=22 measurements from 22 women) were included to estimate the trend of AMH due to natural aging in the tamoxifen-only group. As expected, in regression analysis, we found the patient age to be an important predictor for baseline AMH levels; higher age at study entry was associated with significantly lower AMH levels. This translated into about 11% decrease in serum AMH levels per increasing year of age before the onset of chemotherapy (using baseline data, p<0.0001).

**Table 1.** Characteristics of 142 study participants that remain after the exclusions.

| Variables | Value (%) |
| --- | --- |
| Age | 37.6 ± 4.5 |
| Body mass index | 24.2 ± 4.7 |
| Stage |  |
| (1-2) | 113 (79.5) |
| (3-4) | 27 (19.0) |
| Unknown | 2 (1.4) |
| Estrogen receptor status |  |
| Positive | 123 (86.6) |
| Negative | 19 (13.3) |
| Progesterone receptor status |  |
| Positive | 118 (83.0) |
| Negative | 24 (16.9) |
| HER2 status |  |
| Positive | 31 (21.8) |
| Negative | 111 (78.1) |
| Triple negative | 16 (11.2) |
| BRCA status |  |
| Positive | 14 (9.8) |
| Negative | 76 (53.5) |
| Untested | 52 (36.6) |
| Treatment |  |
| AC-based regimens | 106 (74.6) |
| CMF | 19 (13.3) |
| Tamoxifen-only | 17 (11.9) |
| Tamoxifen use following chemotherapy |  |
| Yes | 97 (77.6) |
| No | 28 (19.7) |

AC: Anthracycline-cyclophosphamide
CMF: Cyclophosphamide, Methotrexate, 5-FU

**Figure 1.**
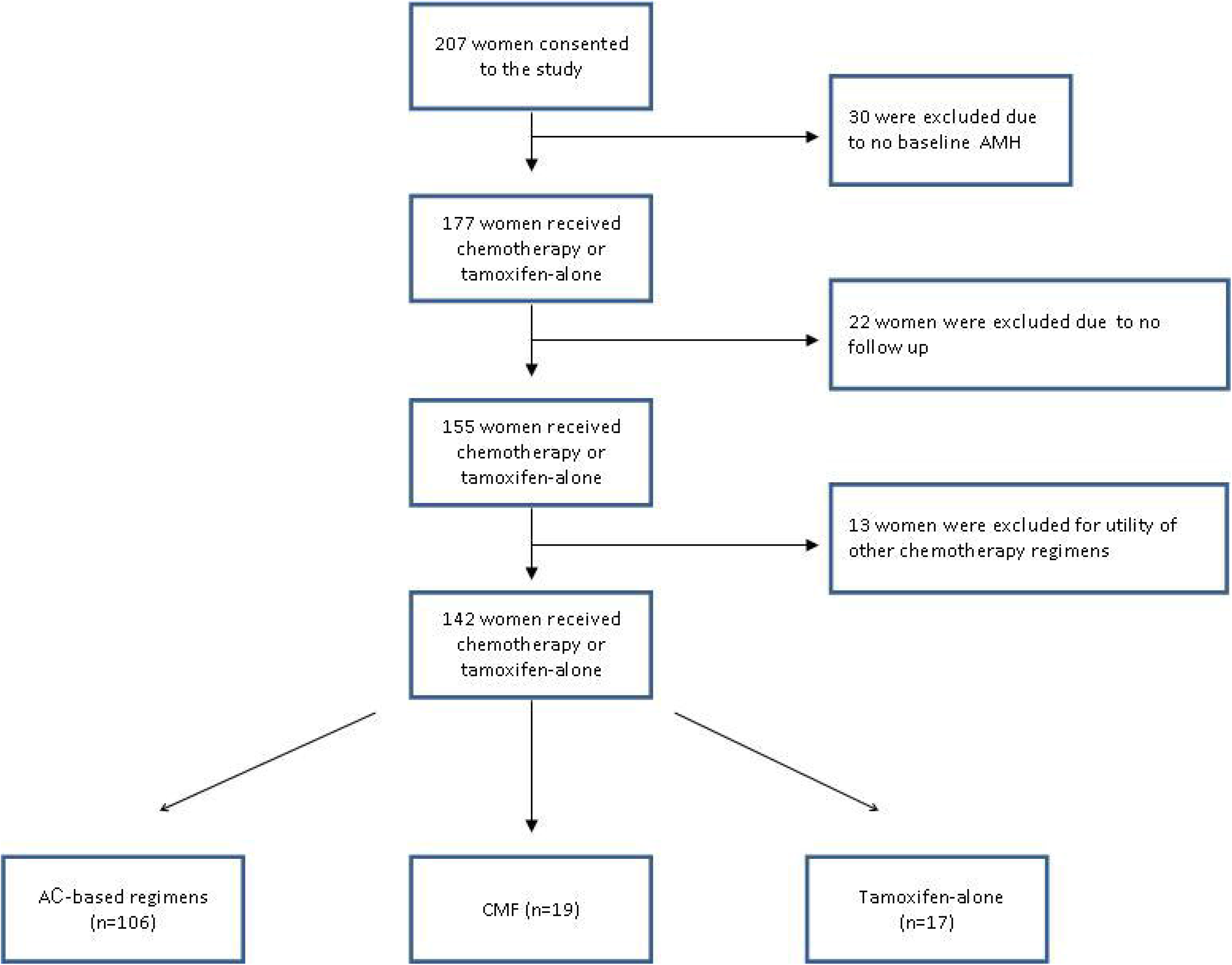
Study inclusion and exclusions flow chart. AC: Anthracycline and cyclophosphamide CMF: Cyclophosphamide, Methotrexate, 5-Fluoro-uracil

### Comparison of serum AMH levels and recovery among the chemotherapy regimens

Mean ages of women receiving AC-based regimens at recruitment were younger than those of women receiving CMF (36.8 ± 4.5 vs. 40.8 ± 3.4 years; p=0.0003 from t-test), respectively. After chemotherapy, AMH levels were sharply lower at the 12-month time point and this decline was similar among the chemotherapy groups, adjusting for baseline age and BMI (Figure 2). The mean serum AMH levels were not significantly different between chemotherapy groups at the 12-, 18- and 24-month time points (p=0.53; Figure 2). In women who received chemotherapy, although the AMH levels continued to recover from 12-up to 24-months post treatment, this recovery was relatively small, and hence, was not clinically meaningful (p=0.97 and 0.04 for 18 and 24 months by signed rank test) (Figure 3). As an example, while the mean/median AMH level was 0.34/0.09 ng/dl at the 12-month time point, it was 0.40/0.06 ng/dl and 0.42/0.07 ng/dl at 18- and 24-month time points for the AC-based regimens group. These values were 0.11/0.03, 0.12/0.02 and 0.20/0.03 at the 3 time points for CMF group. These mean levels are substantially below the threshold for normal ovarian reserve, which is generally 1.1 ng/mL or higher for the age group. AMH levels remained undetectable in 20% (15/76) vs. 38% (5/13) of women in the AC-based regimens vs. CMF groups at 24 months (p=0.16 from Fisher exact test). AMH recovery rate at 18- and 24-months after the completion of chemotherapy was low in AC-based regimens and CMF groups (mean of 10% and 16% at 18 months and 10% and 7% at 24 months, respectively; median <4%) (Figure 2).

**Figure 2.**
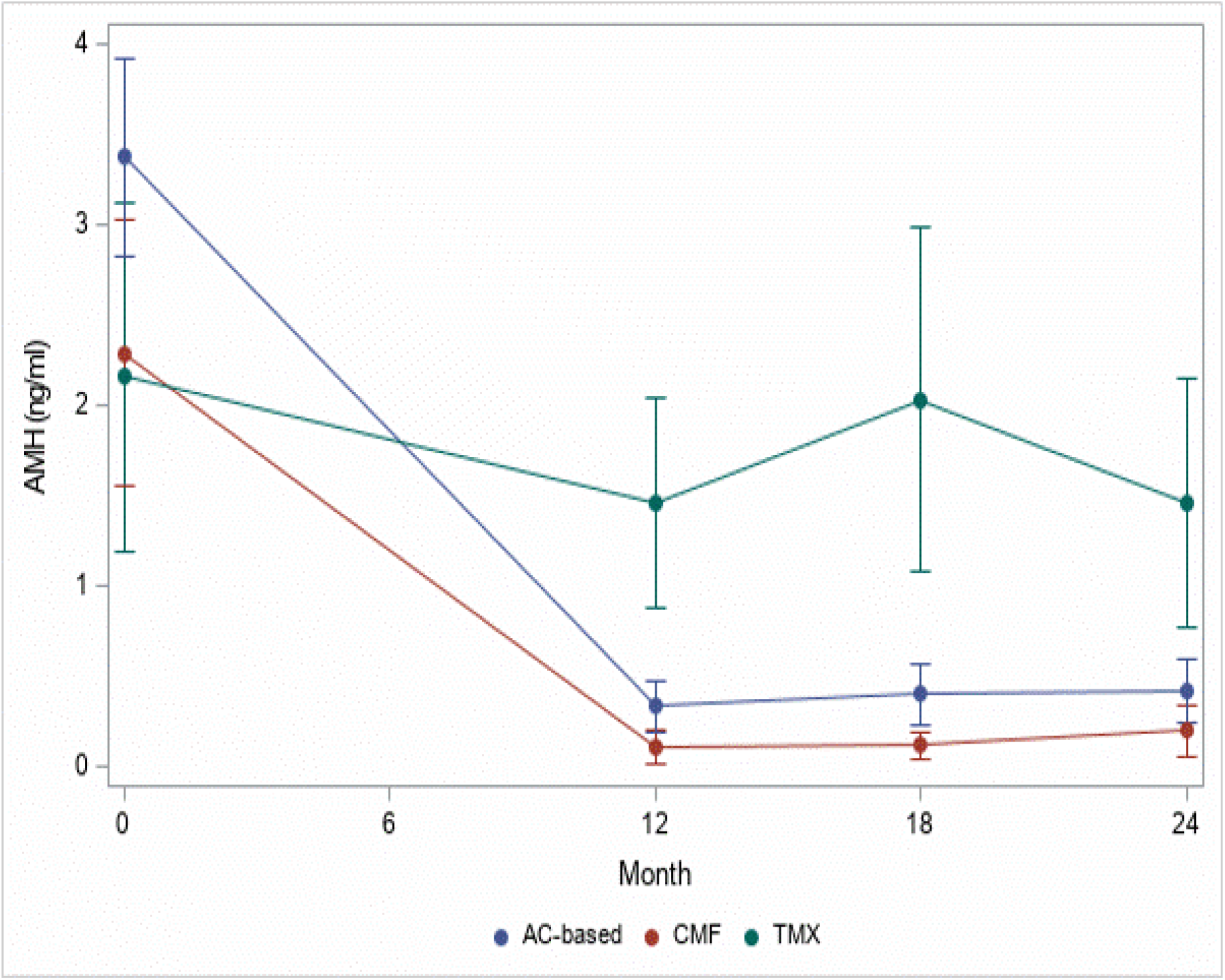
Longitudinal serum AMH changes 12-, 18- and 24-months after the completion of chemotherapy or tamoxifen-only treatment. After a baseline assessment, 142 women were followed up with 12-, 18- and 24-month serum AMH measurements either after the completion of chemotherapy or during tamoxifen-only treatment. Compared to the baseline, there was a significant decline in serum AMH for those who received chemotherapy at all time points but not for tamoxifen-only treatment group. However, serum AMH levels did not differ at any time point among the women who received AC-based or CMF regimens. Women using tamoxifen-alone showed significantly higher AMH levels at 12-, 18- and 24-months (time-trend or slope captured by time*group interaction) compared with women who received AC-based regimens and CMF (both p<0.0001 vs. tamoxifen). Baseline AMH were not statistically different among 3 treatment groups (p=0.45 for df=2). These comparisons indicate that breast cancer chemotherapy regimens result in significant diminishment of ovarian reserve as early as 12 months after the completion of chemotherapy, but tamoxifen does not seem to alter ovarian reserve when given alone beyond natural aging – See Figure S1 for Observed vs. Expected values of AMH in the tamoxifen group. Vertical bar indicates pointwise 95% confidence interval for crude means (i.e., based on raw data), unadjusted for multiplicity. See Table S1 for summary statistics for each subgroup at different time points. AC: Anthracycline and cyclophosphamide; CMF: Cyclophosphamide, Methotrexate, 5-Florouracil; Tmx: Tamoxifen 20 mg/day)

**Figure 3.**
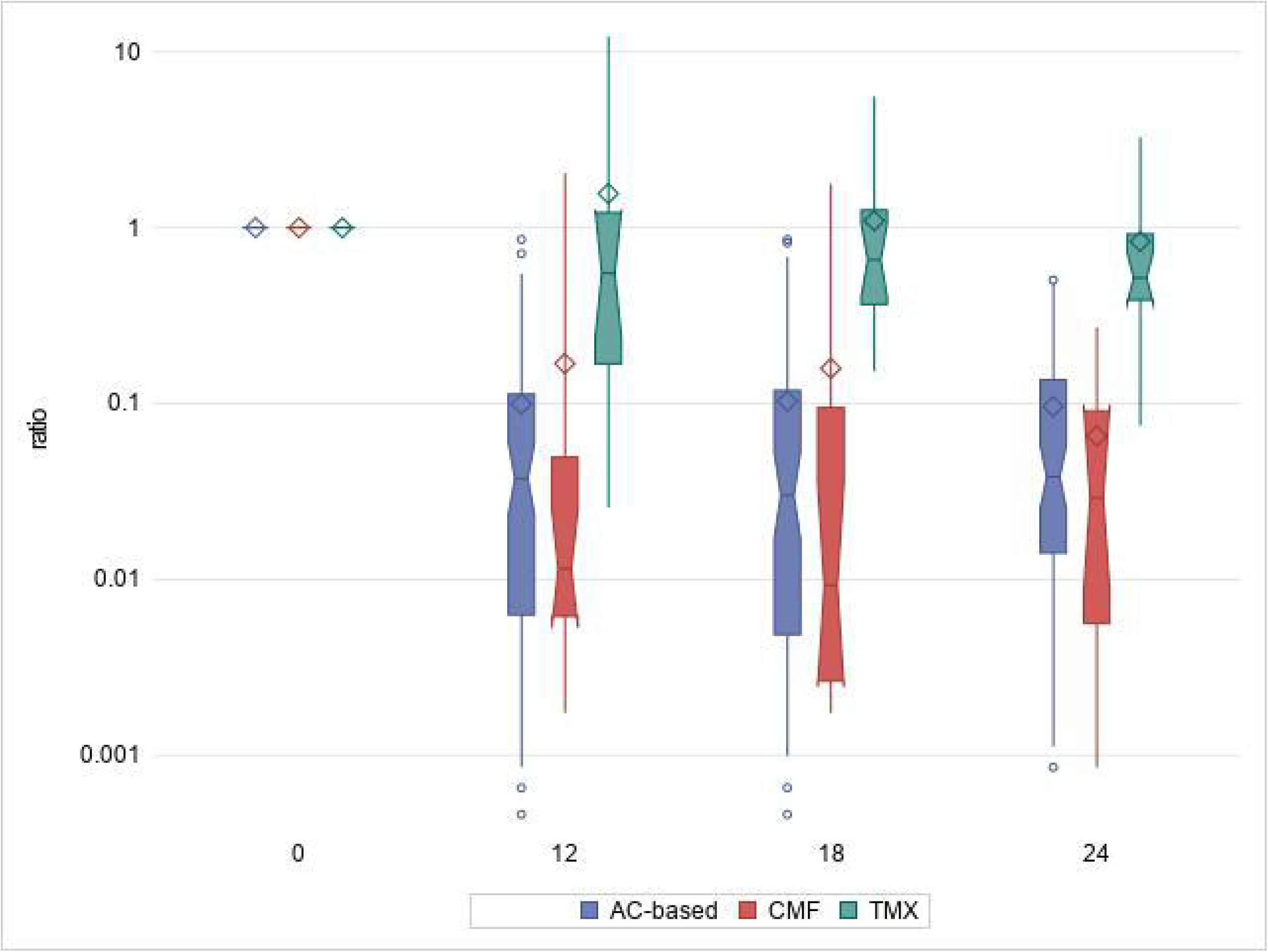
Serum AMH recovery rates in women treated with AC-based, CMF and tamoxifen-only treatments during the 12-, 18- and 24-months follow-up. After a baseline assessment, 142 women were followed up for 24-months with serum AMH measurements either after the completion of chemotherapy or during tamoxifen-only treatment. AMH recovery was calculated by the ratio dividing follow up AMH values by the baseline value at each time point, where ratio=1 means full recovery. AMH recovery rate was similar between women who received treatment with AC-based and CMF chemotherapy regimens 12-, 18- and 24-months after the completion of chemotherapy. Tamoxifen treatment alone was associated with higher recovery rates compared with chemotherapy groups at 12-, 18- and 24-months follow-up. See Table S1 for summary statistics for each subgroup at different time points. AC: Anthracycline and cyclophosphamide; CMF: Cyclophosphamide, Methotrexate, 5-Florouracil; Tmx: Tamoxifen 20 mg/day.

The utility of adjuvant tamoxifen treatment did not affect AMH recovery in AC-based regimens. Likewise, HER-2/neu status was also not influential on the outcomes of AC-based regimens. However, we were not able to analyze the effect of adjuvant tamoxifen treatment or HER-2/neu status on women who received CMF due to the small number receiving either.

We previously showed that women with BRCA mutations may be more prone to losing ovarian reserve, due to oocyte DNA repair deficiency (7). Of the study population, 9.8% carried BRCA mutations, and all were in the AC-based treatment group, with the exception of one. Within this limited sample size, the percentages of women with BRCA mutations were 12.3% (13/106) and 5.3% (1/19) in AC-based regimens and CMF groups, respectively.

### Impact of tamoxifen-only treatment on serum AMH

Seventeen women who received tamoxifen therapy alone had both a baseline and a follow up serum AMH assessment. The mean age of women who received tamoxifen was 40.1 ± 3.3 at the time of enrollment (n=17). After adjustment for age and BMI at the time of assessment, there was an overall small, but significant decline in serum AMH levels during follow-up, compared to the baseline levels in the tamoxifen group (p=0.03) (Figure 2 and 3). However, this decline is well explained by the expected age-induced decline in AMH levels (p=0.79 for comparing cross-sectional vs. longitudinal effects of age; see S Fig for Observed vs. Expected age trend) (31). Women using tamoxifen alone had significantly higher AMH levels (measure by slope or time*treatment group interaction) over 24-months compared with women who received chemotherapy (p<0.0001) (Figure 2).

## Discussion

We evaluated the impact of breast cancer chemotherapy and tamoxifen-only treatment on serum AMH levels for a two-year period. We found that there was a significant decrease in serum AMH regardless of the type of the chemotherapy regimen, but the tamoxifen-only treatment did not affect ovarian reserve. Furthermore, 20% of women in the AC-based regimens and 38% in the CMF group had undetected AMH levels at the end of the two-year follow up period. In contrast, serum AMH levels did not change with tamoxifen treatment during the 24-month follow up beyond natural aging.

One of the key novel findings that our study supplies is the lack of changes in serum AMH levels when tamoxifen was given alone. No other study has investigated serum AMH levels during tamoxifen-only treatment in women who have not received chemotherapy. Likewise, we also did not find any impact of tamoxifen on serum AMH when given subsequent to adjuvant AC-based chemotherapy. While our study is novel with respect to the findings in women receiving tamoxifen-only treatment and its longitudinal design, the impact of tamoxifen treatment on ovarian reserve was evaluated subsequent to chemotherapy in two cross-sectional studies with contrasting results (32,33). One study reported lower serum AMH levels in breast cancer survivors who received adjuvant tamoxifen treatment after chemotherapy, but this conclusion was based on a small sample size (n=10) (32). There was also no adjustment for age as well as the duration of tamoxifen treatment. Another study, in agreement with our findings, reported that post-chemotherapy tamoxifen-users did not have lower ovarian reserve than the tamoxifen-nonusers (33). In that cross-sectional study, 45 survivors had or were taking tamoxifen, while 63 survivors had not received tamoxifen. After adjusting for age, type and duration of chemotherapy exposure, cancer stage, GnRH agonist use and race, the estimated mean AMH was slightly higher for tamoxifen users.

From the pharmacological point of view, the lack of negative impact on ovarian reserve by tamoxifen is not surprising. Because tamoxifen is not a cytotoxic drug, it is not expected to damage primordial follicle reserve. Tamoxifen is an ovarian stimulant and is sometimes used for ovulation induction in anovulatory patients or for IVF treatments for women with breast cancer (34). Since the main production source of AMH is mid-size antral follicles and because tamoxifen alters follicle growth dynamics by stimulation, in theory, tamoxifen may spuriously alter and even increase serum AMH levels without affecting primordial follicle reserve. In our study, after accounting for the age-related decline, we did not find a significant change in serum AMH with 24 months of follow-up. This suggests that any impact of tamoxifen on growing ovarian follicle populations is compensated during chronic treatment. In conclusion, our novel finding in the tamoxifen-only treatment group, as well as the findings on those who received tamoxifen following chemotherapy, assures us that AMH can be reliably used to monitor ovarian reserve in women who or on long-term tamoxifen treatment.

Our study is one of the few studies that provides longitudinal assessment of ovarian reserve and the only one that assessed chemotherapy-induced ovarian reserve decline at multiple time points during a 24-month post-chemotherapy period. A study by Yu et al. prospectively analyzed changes in Mullerian Inhibiting Substance levels (MIS, former nomenclature for AMH) in 26 women with breast cancer using an earlier less sensitive assay than the one we utilized in our study (35). While there were multiple time points, the follow up was only for one year from the initiation of chemotherapy and the patients were not stratified based on treatment regimens (35). The study concluded that breast cancer chemotherapy sharply reduced MIS levels and there was no significant recovery 52 weeks from the initiation or approximately 6-months post-completion of chemotherapy. A later longitudinal study evaluated reproductive function after chemotherapy in 50 premenopausal women with breast cancer, but like the study by Yu et al, patients were only followed for a total of one year including the time spent in treatment (27). In that study, serum AMH levels were assessed at baseline and every three months for a year during and following chemotherapy. The authors found that there was a rapid decline in serum AMH levels three months after the initiation of chemotherapy without a significant recovery during the one-year follow up. In another longitudinal study, AMH recovery was evaluated by sampling before, and 4-weeks and 24-months after the initiation of adjuvant chemotherapy but >50% of those enrolled were on ovarian suppression (36). Among the 101 women, they found a high rate of ovarian reserve impairment at the 24-month follow up. AMH recovery rates were not measured at any mid time points and women were not stratified based on chemotherapy regimens or adjusted for hormonal suppression. In addition to its larger sample size (127 women had at least a 12-month post-treatment assessment), the novelty of our study is that it provides the longest term follow-up with multiple time points up to 24 months (12-, 18- and 24-month assessment in all 80 women) after the completion of chemotherapy, in addition to the comparison of adjuvant chemotherapy types and the tamoxifen-only treatment.

Our study found that serum AMH levels did not meaningfully recover from 12 to 24 months post-chemotherapy (i.e. increase of <0.11 ng/ml in mean and nearly 0 in median). Mean serum AMH levels remained well below normal range at all three time points and indicated severely diminished ovarian reserve. These findings suggest that ovarian reserve assessment as early as one year after the completion of chemotherapy should reflect the final ovarian damage incurred. These findings are also consistent with ovarian physiology as it may take 6 months or longer for the surviving primordial follicles to initiate growth and result in developing follicles that can produce AMH again (37).

Previous studies based on amenorrhea rates suggested that the CMF may be more gonadotoxic than AC-based regimens (38). Our study is the first to compare these protocols with a reliable ovarian reserve marker and did not detect a difference between the AMH recovery patterns of these two chemotherapy regimens. Cyclophosphamide is the most gonadotoxic drug, inducing massive DNA double strand breaks and apoptosis in primordial follicles (4,39). Though CMF delivers a larger cumulative dose of cyclophosphamide compared to AC-based regimens, the latter includes doxorubicin, which is also gonadotoxic. We have shown in human ovarian tissue xenograft experiments that doxorubicin also induces DNA double strand breaks and triggers apoptotic death of primordial follicles similar to cyclophosphamide (4,39). The inclusion of two gonadotoxic drugs in the AC-based regimen may explain its gonadotoxic equivalency to CMF, despite the utility of larger cyclophosphamide doses in the latter. However, we remain reserved with this conclusion as the sample size was smaller for the CMF group and the percentage of women with undetectable AMH were higher with the latter. Moreover, all but one of the women with BRCA mutations were in the AC-based group, which we previously showed to associate with lower AMH recovery (7).

Since the main mechanism of chemotherapy-induced damage to ovarian reserve is via the induction of DNA double strand breaks in primordial follicles and intact BRCA function is required to repair such breaks (5), we have previously hypothesized and shown that women with BRCA mutations experience sharper declines of ovarian reserve after breast cancer chemotherapy (7). We found, based on the same patient population studied in the current report, women with BRCA mutations who received chemotherapy had 3-fold lower AMH recovery compared to those BRCA-mutation-negative or untested during the 12-24 month follow up. These findings were confirmed in a mouse oocyte BRCA knock-out model, where knock-down of BRCA function resulted in a significant increase in oocyte death after *in vitro* exposure to doxorubicin (7). These findings, when used collectively with the current study, provide valuable information for fertility preservation counselling before breast cancer treatment in women with and without BRCA mutations.

Despite its novelty and numerous strengths, our study also had some limitations. We did not know the smoking status of approximately 30% of participants and smoking can result in lower serum AMH levels (40). Though we could not adjust our analysis for smoking status due to the missing information, the incidence of smoking was only 20% in the remaining 70% of the population studied. Given this low incidence and the fact that the study longitudinally compared AMH changes in reference to subjects’ own baseline AMH, smoking status is unlikely to affect our results. We also did not screen the study population for polycystic ovarian syndrome, a disease which could be associated with spuriously elevated AMH levels. However, since we excluded women with irregular periods or amenorrhea from participation and since AMH recovery is permuted relative to each subject’s baseline measurements, it is unlikely that PCOS-screening could have altered our results. Finally, CMF and tamoxifen groups had relatively low sample sizes.

In summation, our study is the first to assess ovarian reserve changes with serum AMH in a prospective longitudinal fashion, with multiple time points up to 24-month post-completion of chemotherapy and in comparison with tamoxifen-only treatments. It shows that the chemotherapy regimens used for breast cancer treatment are highly damaging to ovarian follicle reserve, and tamoxifen-only treatment does not affect serum AMH assessment. However, larger studies may be needed to determine the precise differences of gonadotoxicity between the specific chemotherapy regimens. In the meantime, our study provides novel information to use when counselling young women with breast cancer before they receive adjuvant chemotherapy and tamoxifen-only treatments.

## Data Availability

All data referred to in the manuscript is available if corresponding author approves.

## Contributions

Conception of the idea: KO; Design: KO, MD, SG, SP; Study execution: SG, MD, KO, GB, VT, ET; Provision of study materials: KO, MD, SG, TC; Manuscript writing: KO, VT, SG, GB, HB; Statistical Analysis: VT, HB; Final approval: All authors.

## Acknowledgments

We thank Ansh Laboratories for their assistance in the analysis of serum AMH levels and Angelena Crown, MD for help with data extraction.

## Supplements

**Figure S1.** Observed vs. Expected values of AMH for tamoxifen group, where expected AMH value was estimated as a function of age using baseline samples (n=22). Using mean (left) and median (right).

**Table S1.** Summary statistics for AMH for 3 treatment groups and 4 time points.

| Month | Treatment | N | Mean | SD | Median | Lower<br>Quartile | Upper<br>Quartile |
| --- | --- | --- | --- | --- | --- | --- | --- |
| 0 | AC-based | 106 | 3.37 | 2.89 | 2.69 | 1.28 | 4.49 |
|  | CMF | 19 | 2.29 | 1.64 | 1.81 | 0.89 | 3.87 |
|  | TMX | 22 | 2.01 | 1.99 | 1.43 | 0.38 | 3.01 |
| 12 | AC-based | 95 | 0.34 | 0.71 | 0.09 | 0.02 | 0.32 |
|  | CMF | 18 | 0.11 | 0.22 | 0.03 | 0.003 | 0.13 |
|  | TMX | 14 | 1.47 | 1.11 | 1.6 | 0.50 | 2.50 |
| 18 | AC-based | 82 | 0.40 | 0.80 | 0.06 | 0.005 | 0.34 |
|  | CMF | 18 | 0.12 | 0.16 | 0.02 | 0.003 | 0.24 |
|  | TMX | 16 | 2.03 | 1.94 | 1.66 | 0.38 | 3.01 |
| 24 | AC-based | 76 | 0.42 | 0.77 | 0.07 | 0.01 | 0.49 |
|  | CMF | 13 | 0.20 | 0.26 | 0.03 | 0.003 | 0.43 |
|  | TMX | 16 | 1.46 | 1.41 | 0.92 | 0.23 | 2.47 |

AC-based: Anthracycline and cyclophosphamide based treatment
CMF: Cyclophosphamide, Methotrexate, 5-Fluorouracil
TMX: Tamoxifen

## References

1. Héry C, Ferlay J, Boniol M, Autier P. Changes in breast cancer incidence and mortality in middle-aged and elderly women in 28 countries with Caucasian majority populations. Ann Oncol 2008;19:1009–18

2. Bray F, Ferlay J, Soerjomataram I, Siegel RL, Torre LA, Jemal A. Global cancer statistics 2018: GLOBOCAN estimates of incidence and mortality worldwide for 36 cancers in 185 countries. CA Cancer J Clin 2018;68:394–424

3. Bedoschi G, Navarro PA, Oktay K: Chemotherapy-induced damage to ovary: Mechanisms and clinical impact. Futur Oncol, 2016;12:2333–44

4. Oktem O, Oktay K: Quantitative assessment of the impact of chemotherapy on ovarian follicle reserve and stromal function. Cancer 2007;110:2222–2229

5. Soleimani R, Heytens E, Darzynkiewicz Z, Oktay K. Mechanisms of chemotherapy-induced human ovarian aging: double strand DNA breaks and microvascular compromise. Aging (Albany NY) 2011;3:782–793

6. Tham Y-L, Sexton K, Weiss H, Elledge R, Friedman LC, Kramer R. The Rates of Chemotherapy-Induced Amenorrhea in Patients Treated With Adjuvant Doxorubicin and Cyclophosphamide Followed by a Taxane. Am J Clin Oncol 2007;30:126–132

7. Oktay KH, Bedoschi G, Goldfarb SB, Taylan E, Titus S, Palomaki GE et al: Increased chemotherapy-induced ovarian reserve loss in women with germline BRCA mutations due to oocyte deoxyribonucleic acid double strand break repair deficiency. Fertil Steril 2020;113:1251–1260.

8. Turan V, Oktay K: BRCA-related ATM-mediated DNA double-strand break repair and ovarian aging. Hum Reprod Update 2020;26:43–57

9. Dunlop CE, Anderson RA: Uses of anti-Mullerian hormone (AMH) measurement before and after cancer treatment in women. Maturitas 2015;80:245–250

10. Durlinger ALL, Visser JA, Themmen APN: Regulation of ovarian function: The role of anti-Mullerian hormone. Reproduction 2002;124:601–9

11. Fleming R, Seifer DB, Frattarelli JL, Ruman J. Assessing ovarian response: antral follicle count versus anti-Mullerian hormone. Reprod Biomed Online 2015;31:486–96

12. Hansen KR, Hodnett GM, Knowlton N, Craig LB. Correlation of ovarian reserve tests with histologically determined primordial follicle number. Fertil Steril 2011;95:170–5

13. Iliodromiti S, Anderson RA, Nelson SM. Technical and performance characteristics of anti-Mullerian hormone and antral follicle count as biomarkers of ovarian response. Hum Reprod Update 2015;21:698–710

14. Van Rooij IAJ, Broekmans FJM, Scheffer GJ, Looman CWN, Habbema JF, de Jong FH, et al: Serum antiMullerian hormone levels best reflect the reproductive decline with age in normal women with proven fertility: A longitudinal study. Fertil Steril 2005;83:979–87

15. Bentzen JG, Forman JL, Johannsen TH, Pinborg A, Larsen EC, Andersen AN et al: Ovarian antral follicle subclasses and anti-Mullerian hormone during normal reproductive aging. J Clin Endocrinol Metab 2013;98:1602–11

16. Jeppesen J V., Anderson RA, Kelsey TW, Christiansen SL, Kristensen SG, Jayaprakasan K et al: Which follicles make the most anti-Mü llerian hormone in humans? Evidence for an abrupt decline in AMH production at the time of follicle selection. Mol Hum Reprod 2013;19:519–27

17. Durlinger ALL, Gruijters MJG, Kramer P, Karels B, Kumar TR, Matzuk MM et al: Anti-Mullerian hormone attenuates the effects of FSH on follicle development in the mouse ovary. Endocrinology 2001;142: 4891–9

18. Durlinger ALL, Gruijters MJG, Kramer P, Karels B, Ingraham HA, Nachtigal MW et al: Anti-Mullerian hormone inhibits initiation of primordial follicle growth in the mouse ovary. Endocrinology 2002;140:5789–96,

19. Dewailly D, Andersen CY, Balen A, Broekmans F, Dilaver N, Fanchin R et al: The physiology and clinical utility of anti-Mullerian hormone in women. Hum Reprod Update, 2014;20:370–85

20. Riggs R, Kimble T, Oehninger S, Bocca S, Zhao Y, Leader B et al: Anti-Mullerian hormone serum levels predict response to controlled ovarian hyperstimulation but not embryo quality or pregnancy outcome in oocyte donation. Fertil Steril 2011;95:410–2

21. Riggs RM, Duran EH, Baker MW, Kimble TD, Hobeika E, Yin L et al: Assessment of ovarian reserve with anti-Mullerian hormone: a comparison of the predictive value of anti-Mullerian hormone, follicle-stimulating hormone, inhibin B, and age. Am J Obstet Gynecol 2008;199:202 e1–8

22. Broer SL, Eijkemans MJC, Scheffer GJ, van Rooij IA, de Vet A, Themmen AP et al: Anti-Mullerian hormone predicts menopause: A long-term follow-up study in normoovulatory women. J Clin Endocrinol Metab. 2011;96:2532–9

23. Freeman EW, Sammel MD, Lin H, Gracia CR. Anti-Mullerian hormone as a predictor of time to menopause in late reproductive age women. J Clin Endocrinol Metab 2012;97:1673–80

24. Tehrani FR, Solaymani-Dodaran M, Tohidi M, Gohari MR, Azizi F. Modeling Age at Menopause Using Serum Concentration of Anti-Mullerian Hormone. J Clin Endocrinol Metab 2013;98:729–735

25. Anderson RA, Cameron DA. Pretreatment serum anti-Mullerian hormone predicts long-term ovarian function and bone mass after chemotherapy for early breast cancer. J Clin Endocrinol Metab, 2011;96:1336–43

26. Henry NL, Xia R, Schott AF, McConnell D, Banerjee M, Hayes DF. Prediction of Postchemotherapy Ovarian Function Using Markers of Ovarian Reserve. Oncologist 2014;19:68–74

27. Anderson RA, Themmen APN, Al-Qahtani A, Groome NP, Cameron DA. The effects of chemotherapy and long-term gonadotrophin suppression on the ovarian reserve in premenopausal women with breast cancer. Hum Reprod, 2006;21:2583–92

28. Lee S, Ozkavukcu S, Heytens E, Moy F, Oktay K. Value of Early Referral to Fertility Preservation in Young Women with Breast Cancer. J Clin Oncol 2010;28:4683–6

29. Su HI, Sammel MD, Green J, Velders L, Stankiewicz C, Matro J et al: Antimullerian hormone and inhibin B are hormone measures of ovarian function in late reproductive-aged breast cancer survivors. Cancer 2010;116:592–599

30. Oktay K, Harvey BE, Partridge AH, Quinn GP, Reinecke J, Taylor HS et al: Fertility preservation in patients with cancer: ASCO clinical practice guideline update. J Clin Oncol 2018;36:1994–2001

31. G Fitzmaurice, Laird N, Ware J: Longitudinal Analysis. Second edition, Wiley, 2011Available from: https://content.sph.harvard.edu/fitzmaur/ala2e/

32. Partridge AH, Ruddy KJ, Gelber S, Schapira L, Abusief M, Meyer M et al: Ovarian reserve in women who remain premenopausal after chemotherapy for early stage breast cancer. Fertil Steril 2010;94:638–44

33. Shandley LM, Spencer JB, Fothergill A, Mertens AC, Manatunga A, Paplomata E et al: Impact of tamoxifen therapy on fertility in breast cancer survivors. Fertil Steril 2017;107:243–252.e5

34. Oktay K, Buyuk E, Davis O, Yermakova I, Veeck L, Rosenwaks Z. Fertility preservation in breast cancer patients: IVF and embryo cryopreservation after ovarian stimulation with tamoxifen. Hum Reprod 2003;18:90–5

35. Yu B, Douglas N, Ferin MJ, Nakhuda GS, Crew K, Lobo RA et al: Changes in markers of ovarian reserve and endocrine function in young women with breast cancer undergoing adjuvant chemotherapy. Cancer 2010;116: 2099–2105

36. Trapp E, Steidl J, Rack B, Kupka MS, Andergassen U, Jückstock J et al., Anti-Müllerian hormone (AMH) levels in premenopausal breast cancer patients treated with taxane-based adjuvant chemotherapy - A translational research project of the SUCCESS A study. Breast. 2017;35:130–135

37. Oktay K, Newton H, Mullan J, Gosden RG. Development of human primordial follicles to antral stages in SCID/hpg mice stimulated with follicle stimulating hormone. Hum Reprod 1998;13:1133–1138

38. Bines J, Oleske DM, Cobleigh MA. Ovarian function in premenopausal women treated with adjuvant chemotherapy for breast cancer. J Clin Oncol 1996;14:1718–29

39. Li F, Turan V, Lierman S, Cuvelier C, De Sutter P, Oktay K. Sphingosine-1-phosphate prevents chemotherapy-induced human primordial follicle death. Hum Reprod 2014;29:107–13

40. Plante BJ, Cooper GS, Baird DD, Steiner AZ. The impact of smoking on anti-Mullerian hormone levels in women aged 38 to 50 years. Menopause 2010;17:571–6

